# A One Health approach to pastoral (im)mobility, health, and disease: a qualitative participatory study in Plateau State, Nigeria

**DOI:** 10.1101/2023.09.01.23294938

**Authors:** Dorien Hanneke Braam, Muhammad-Bashir Bolajoko, Charlotte Christiane Hammer

**Affiliations:** Disease Dynamics Unit, University of Cambridge, Cambridge, UK; National Veterinary Research Institute, Vom, Plateau State, Nigeria; Disease Dynamics Unit, Department of Veterinary Medicine, University of Cambridge, Madingley Road, CB3 0ES, Cambridge, United Kingdom

**Keywords:** Migration, Nigeria, zoonoses, social determinants of health, conflict

## Abstract

**Background:** In Plateau State, pastoralism has historically been a cost effective and resilient economic system well-suited to the ecological context. However, changes in land use and conflict have increasingly changed patterns of mobility. Pastoralist movement is now often associated with zoonotic disease transmission, environmental degradation and conflict, increasingly resulting in forced sedentarisation. Rather than a direct outcome of population movement however, animal, human and zoonotic disease drivers are complex and influenced by a range of socio-economic and environmental factors. The interlinkages of (im)mobility and health requires better understanding of underlying vulnerabilities to disease, which we aim to address in this study.

**Methods:** Using a multisite case study methodology we investigated pastoralists’ animal and human health concerns and priorities in Plateau State, Nigeria. We deployed participatory tools, including open-ended Focus Group Discussions, transect walks, mapping exercises, calendars and matrices. Data were analysed using a One Health conceptual framework.

**Results:** We interviewed 105 participants, from transhumance, migratory and sedentary households, dependent on livestock and small-scale crops. While transhumance was often preferred, participants had become sedentary as a result of insecurity, loss in livestock, and household characteristics. Humans and animals suffered from several endemic diseases, including zoonoses, however veterinary and human health services are only available in larger towns, and people mostly rely on community (animal) health workers and self-medication. Both transhumance and sedentary livestock keepers face challenges around forage grazing, regularly blocked by landowners, sometimes escalating into conflict.

**Conclusions:** While conflict and changes in land use affected animal and human disease patterns, underlying political, social and economic risk factors were important determinants of health. There is a need for more inclusive, transdisciplinary, multilevel approaches to address animal and human disease, based on better contextualization of the challenges, through the participation of affected communities.

**Impacts:**

- This study uses participatory approaches to investigate the under researched area of mobility and zoonotic disease dynamics.
- We found that animal and human mobility may mitigate or exacerbate zoonotic disease risk, impacting conflict, in turn leading to forced migration.
- Conflict and crime related security challenges need to be addressed simultaneously with mitigating structural inequality, through increasing pastoral social and economic capital, increase access to grazing routes and water, secure land tenure arrangements, and the provision of (veterinary) public health to reduce zoonotic disease risks in the region.

## Introduction

Nigeria is Africa’s most populous country, with a population of over 200 million people (FEWSNET, 2018). Agriculture contributes about 21.2 percent to Gross Domestic Product (GDP), with livestock contributing 9 percent (Majekodunmi, 2022), and 36.5 percent to employment (Ihekweazu et al., 2021). The national livestock herd is estimated at 18.4 million cattle, 43.4 million sheep, 76 million goats and 180 million poultry (FAO, 2019). Pastoralism, a subsistence strategy based on the herding of animals, practiced by the majority of ruminant farmers (Majekodunmi, 2022) accounts for 82 percent of cattle. Herd sizes are extremely variable, with most herds in the ranges of 10 to 20 cattle and 50 to 100 cattle (Majekodunmi, Fajinmi, Dongkum, Shaw, & Welburn, 2014). This transhumant production system is based on the seasonal and cyclical migration of livestock, with non-herding members of the community engaging in crop production (IOM, ECOWAS, & ICMPD, 2017). There is a lack of data on pastoralism in sub-humid zones (Majekodunmi et al., 2014), as well as the interlinkages of migration and zoonotic disease dynamics. While rabies, avian influenza, Ebola Virus Disease (EVD), swine influenza and anthrax are considered zoonoses of most concern (Ihekweazu et al, 2021), there is little data available on the actual zoonotic disease burden in the region.

Pastoralists are often held responsible for overgrazing and disease spread (Sani Ibrahim, Ozdeser, Cavusoglu, & Abdullahi Shagali, 2021), even though pastoralism may actually contribute to healthier environments, as continuous movement enables the recovery of natural resources (Manzano et al., 2021). The association with disease transmission, environmental degradation and conflict, increasingly results in forced sedentarisation of pastoral communities (Okello et al, 2014). However, this may increase disease burden, as seasonal vectors such as mosquitoes and tsetse flies can no longer be avoided (Leach et al, 2017). Rather than a direct outcome of population movement therefore, zoonotic disease drivers are complex and influenced by a range of socio-economic and environmental factors (Braam et al, 2021a). The interlinkages of (im)mobility and zoonotic health requires better understanding of underlying vulnerabilities to disease, which we aim to address in this study.

## Materials and Methods

We used a participatory multisite case study methodology to investigate pastoralists’ zoonotic health concerns and priorities in Plateau State, Nigeria. Fieldwork was conducted in five communities during February 2022. We employed participatory methods to enhance participants’ control, implemented by researchers from different disciplines, including public health, veterinary medicine and the social sciences, to ensure interdisciplinarity and strengthen research design and analysis. Open-ended Focus Group Discussions (FGDs) to allow for refocusing of the interview based on participants’ priorities. Findings were analysed and interpreted using a One Health conceptual framework, which allows for a comprehensive consideration of human, animal and environmental health, taking into account complex historical, political, and socio-economic dynamics.

### Study area and participants

Historically, the central plain of Plateau State was largely uninhabited, open savannah woodland, which offered grazing areas and the absence of tsetse and mosquitoes to the hill tribes resettled there by the British colonizers (Majekodunmi et al., 2014). Fulani established themselves across the Plateau and on the lowland plains, for its fertile environment and relatively low levels of disease, adopting a largely agro-pastoralist lifestyle around one hundred years ago (Higazi, 2016). Fulani are the primary pastoral population in Nigeria, a diverse group numbering around 15.3 million people. In addition to livestock keeping, most Fulani grow crops such as maize, vegetables and millet on small pieces of land up to 10 acres, a way to spread risk in response to increased competition over natural resources (Majekodunmi, Dongkum, Langs, Shaw, & Welburn, 2017; Majekodunmi et al., 2014). The Fulani are still considered as ‘settlers’, while the ‘indigenes’ Christian tribes retain an advantage in terms of social, political and economic benefits (Majekodunmi et al., 2014). This has caused tensions which spilled over into violent conflict in 2001 and have flared up regularly since. In rural areas, conflicts focus primarily on livestock grazing and crop encroachment.

The Fulani communities interviewed for this study include transhumance, migratory and sedentary households. Community leaders and 10-12 households were selected using a purposive sampling method based on mobility and livelihood, to partake in Focus Group Discussions (FGDs). We interviewed 105 participants, 53 women and 52 men, of whom 29 practices transhumance while 23 remained sedentary. All participant households owned mixed herds primarily for subsistence, and supported their livelihoods through crop farming, and labour migration, while in one community (MA4) some participants earned a livelihood by providing transportation. [sentence deleted for anonymization].

### Data collection and analysis

Fieldwork was conducted in Fulfulde language by six researchers from the Hausa (5) and Fulani (1) ethnic groups, which included experienced veterinary staff, community animal health workers (CAHW) and an existing network of pastoralist community leaders. The semi-structured FGDs included questions on demographic and socio-economic community and household characteristics, individual and community mobility and migration experiences, human and animal health, and health-seeking practices. The interview guides are available as supplementary material to this manuscript. Interviews were recorded with the participants’ consent, and transcripts and fieldwork notes translated into English for analysis. Quotes and findings presented in the Results section were anonymised through assigning interview codes which represent the gender of those interviewed during the FGD, and the geographical location where the FGD was held. Observations within and around the household and community were recorded using field notes and photos and analysed to contextualize and triangulate interview responses. Data analysis was conducted manually in English, using a thematical analysis approach (Attride-Stirling, 2001). Study findings were interpreted using concepts from the One Health framework approach to analyse the interlinkages of human, animal and environmental health and to position the findings within current trends and developments in Plateau State and Nigeria.

## Results

### Communities and their environment

Besides keeping ruminants and poultry for milk, manure and meat, all participants planted crops for food and income. Most crop produce was destined for home consumption, while leftovers were used for animal feed or sold for extra income or school fees (WM1). Participants’ primary concern was the reduction in access to land for subsistence crop farming and grazing (WM1; WK3). While livestock could feed on crop leftovers during dry season (MK3), in rainy season grazing areas were blocked by farmers cultivating their land (MK3; ML5). Elsewhere, government farming occupied former pasture (ML5). Another concern was the limited water supply, in particular during the dry season. Water was obtained from different sources: central boreholes (MM1; WS2), private wells (MM1), constructed or natural dams (WM1; WS2), and/ or water trucking (WA4). None of these sources guaranteed sufficient water supply, however. Over the years, dry season farming has taken over communal water sources (MM1). Some participants had made (paid) arrangements with farmers (MM1), whereby their animals were allowed to drink water from irrigation canals or streams on farmland (WM1; ML5). Other communities had invested in generators, pumping machines and water basins, however strained resources limited the water supply to specific hours, with people and herds taking turns (WM1). Elsewhere, the closest water source was as far away as two hours travel (WK3). During rainy season in June and July (WM1) rivers filled gullies with water for the animals, posing an erosion risk (MS2). Observations confirmed that erosion was present in four out of five communities, causing occasional rockfall during rainy season (WA4). Other environmental degradation was caused by deforestation (MA4), with 64 percent of the land at risk (FAO, 2019).

Even though the sale of animals was an important part of participants’ livelihood, there were few animal markets in the region, with participants travelling up to two hours (WA4). Travelling to and from the markets was not without its risks from thieves and other insecurity issues (MM1). Pretend ‘security agents’ at illegal checkpoints sometimes required bribes (MM1), while property theft included stealing motorcycles and small animals (MA4; ML5). More significant theft occured at night when men returned from the markets with their profits (WA4; ML5). Road accidents were common, particularly during rainy season (MK3; ML5). Only in one community none of the participants had faced security challenges on their way to the market (WL5).

### (Im)mobility

Communities consisted of a combination of transhumant and settled/ sedentary households. Transhumance - moving livestock between grazing grounds in a seasonal cycle – was conducted by men and considered necessary to find good quality pastures for the animals (MM1; MS2), and/ or water sources (MK3). Some herd owners hired boys from Langtang North and South to help with grazing, paid in-kind after the season (WS2; WA4). Transhumance had become increasingly challenging however, with grazing routes and seasons changing as a result of insecurity related to conflict and crime (WM1). While generally migration for grazing occurred during the dry season, starting in December/ January and returning between April - June, with varied timing depending on the origin and destination locations, one key informant mentioned that cattle left for grazing in November and returned in August (WM1). Some participants had experienced cattle rustling (MS2; MA4), while others lost sheep and goats to theft (MK3), with one community losing some just the day before the interview (WK3). Sometimes children and livestock were attacked during grazing (WS2). During the worst farmer - herder clashes, some children were even killed (WM1), although participants noted that in some communities, relationships improved in the past few years (MM1; WS4).

Cattle migration routes were increasingly encroached by crop farmers (WS2) or urbanization (ML5; MA4). Crop farming encroachment was a particular risk during the rainy season, when farmers planted their crops near or on the migratory routes (WL5), and respondents were careful not to enter farmlands to prevent conflict (MK3). Some had decided to graze their animals along the sides of the road to avoid clashes (WL5), while moving at night to minimize the risk of road traffic accidents (MM1). Fishing and irrigation farming had taken over migratory watering points (ML5).

As a result of the challenges encountered during migration, including conflicts (MM1), lacking safe cattle migration routes (WM1) and blockades, some participants had stopped transhumance altogether. Sedentary participants generally owned fewer animals (MS2; MA4) and faced challenges in feeding these (WS2; WK3). One participant explained how ‘during rainy season there is abundant water, but nowhere to forage, while during the dry season there is no water, but abundant feed’ (WA4). Participants mentioned that for sedentary households, animal rearing is easier in the dry season (WK3), when farmers may allow livestock to feed on crop harvest leftovers (ML5) in return for manure (WS2), or for a payment of up to thirty thousand naira (US$40.4)] (WA4). However, the majority of farmers prevented animal grazing by burning the stalks on their fields immediately after harvesting.

### Health and disease

Most participants considered transhumance healthier for the animals (MM1; MA4), however others noted that even during migration there was not always sufficient water, and diseases in grazing areas affected humans and animals (WM1). Sick animals were kept at home, rather than continuing migration (WK3). Participants were familiar with a range of animal diseases, often known by their local names and/ or symptoms, with the most common diseases named: *huhu* (contagious bovine pleuropneumonia (CBPP), *hanta* (fasciolosis), *boru* (Foot and Mouth Disease (FMD), *saifa* (Anthrax), and *samore* (trypanosomiasis) in cattle; *mura* (Peste des Petits Ruminants (PPR), *dingishi* (foot rot), ecto- and endoparasites in sheep and goats; and Newcastle Disease (NCD) in poultry, reflecting findings from the literature (Ndahi et al, 2012). Table 1 gives an overview of the occurance of reporting of the most reported diseases across the fieldwork communities. Besides disease, animals and humans were also at risk of wild animals, including *dilah* (fox, Vulpes vulpes), and *tunku* (wildcat, Felis silvestris) (MS2) and hyenas (MK3), while scorpion- and snake bites occured mainly during the rainy season (WL5)/ March to June (WK3). In response to wild animals and other threats, all participants kept dogs to guard their property. Most participants were familiar with the risk of *haukan kere* (rabies) from dogs (MA4; ML5), however not all dogs were vaccinated, instead they were euthanized when sick (WK3; WL5). Inside the house, participants kept cats to control rodents (MM1). Cats were allowed to sleep on the bed (WM1) and were considered healthy: ‘cats don’t fall sick’ (WM1; WS2), although in one community people understood from the veterinarian that cats may transmit respiratory diseases (WA4; MA4).

**Table 1:** most reported livestock diseases per community.

| Livestock disease reported | CBPP | Fasciolosis | FMD | Anthrax | Trypanosomiasis | PPR | Foot rot | Parasites | NCD |
| --- | --- | --- | --- | --- | --- | --- | --- | --- | --- |
| MM1 | X | X | X |  |  | X | X |  |  |
| WM1 | X | X | X | X |  |  |  | X |  |
| WS2 | X | X | X |  | X | X |  |  | X |
| MS2 | X | X | X |  | X | X | X |  | X |
| WK3 | X | X | X | X |  | X | X |  | X |
| MK3 | X | X | X | X | X | X | X | X | X |
| WA4 | X | X | X |  | X | X |  | X | X |
| MA4 | X | X | X |  | X | X | X | X | X |
| ML5 | X | X | X | X | X | X | X | X |  |

If ruminants fell ill, traditional treatments were used, while some participants called a veterinary officer, although qualified staff and equipment were not always available (MM1). CAHW sometimes visited the villages to treat and vaccinate animals on demand (MS2; WK3; MK3), while veterinary advice was also obtained at veterinary clinics and dispensaries in larger towns and cities (MA4), up to 3.5 hours away (WA4). Drugs were often self-administered (MK3; ML5), and the use of long-acting antibiotics was common (MK3). In the past, quacks treated some animals with fake drugs, ‘costing their husbands over 300 thousand naira’ (US$404) (WS2).

## Discussion

### Structural inequalities

We found that transhumance mobility decisions depended on people’s cattle herd size, considerations around personal safety and security, household circumstances and past experiences. Conflict and related insecurity influence migration patterns (Sani Ibrahim et al., 2021), however rather than a driver of displacement, we find that insecurity is a reason to become sedentary. The loss of livestock and the need for income diversification are important household determinants of migration dynamics (Sani Ibrahim et al., 2021). Those pastoralists fearing the loss of their livelihoods consider sedentarisation or migration, with direct implications to global health and security, related to the competition for land and natural resources between farmers and pastoralists (Sani Ibrahim et al., 2021). Sedentarisation also increases risks of erosion, through the constant use of water points and gullies, and overgrazing, as well as infectious disease amid increased population densities. As a result of climate change and environmental degradation, migration is likely to increase (Olaniyan & Okeke-Uzodike, 2015), as the pastoralist livelihood is challenged (SIPRI, 2022).

Attitudes to pastoralism in Nigeria are mixed. To those in favour, transhumant pastoralism is seen as a highly productive and resilient economic system, and an effective and appropriate way to use grazing resources (Ducrotoy et al., 2018), well adapted to the ecological context of West Africa (IOM et al., 2017). By opponents of transhumance, pastoral livestock production is considered increasingly unsustainable as a primary source of livelihood due to the changing socio-economic context, climatic conditions, and rapid population growth (Sani Ibrahim et al., 2021). In response to the growing population and limitations to available land, the government introduced a 10-year National Livestock Transformation Plan (NLTP) in 2019, introducing livestock identification, hygiene and welfare compliant slaughter measures, linking initiatives for rehabilitation of existing grazing reserves, pasture management and dairy collection centres, with the ultimate aim to limit pastoral mobility (Majekodunmi, 2022). This renewed proposal for ranching to stop pastoralist transhumance and increase intensification, ignores the political and economic factors which caused previous attempts at implementing grazing reserve systems to fail (UNOWAS, 2018), creating distrust and increased opposition from pastoralists (ICC, 2021).

### Pathways to disease

Our results indicate that increased sedentarisation and marginalisation of pastoralists affect livelihoods, but also animal, human and environmental health and disease dynamics. While most transhumance households migrate for forage, others migrate for (animal) health purposes, although some reported that animal health and production levels are not necessarily better in those who move. Where grazing is optimal, nutrition may improve animals’ health status, however incursion into new environments and population density leads to increased vector contact and disease transmission (Majekodunmi, 2022). Meanwhile, when moving to urban centres, pastoralists will lose livestock and nutrition, reducing their exposure to zoonoses, as they no longer live with their animals. These complex dynamics are reflected in the literature, highlighting a gap in knowledge as to how (changes in) mobility affect animal and human health (Braam et al, 2021b).

Both transhumance and sedentary livestock keepers reported grazing challenges, which is regularly blocked by landowners’ farming practices. This creates pathways to disease, for instance, moving away from the tsetse flies during the rainy season might no longer be possible (Ducrotoy et al., 2018; Okello, Majekodunmi, Malala, Welburn, & Smith, 2014). A number of diseases are endemic and present year-round or seasonal in both humans and animals. Participants were aware of a range of infectious and zoonotic disease risks, including drinking from standing water and dog bites. Responses to disease vary however, based on the availability of veterinary and health services. These are often only available in larger towns, and people mostly rely on CAHWs and self-medication. Migration changes the distance to markets and services, already suboptimal in origin villages. A 2005 survey of nomadic Fulani found that 99 percent of children had not received routine childhood immunisations due to their mobility and remote locations (Wild et al., 2020).

Faced with the high burden of infectious diseases, including zoonoses in pastoral areas, in December 2019 the government launched a national One Health strategic plan to integrate human, animal and environmental health management. A high-level disease prioritization workshop defined rabies, avian influenza, EVD, swine influenza and anthrax as national priority zoonoses, primarily focusing on diseases impacting a country’s trade and economy, rather than the concerns of smallholders and poor rural livestock keepers (Ihekweazu et al., 2021). Our participants considered neglected endemic diseases as most impactful instead, highlighting the importance for inclusion of all stakeholders in these exercises.

### Safety first

In our study, structural inequalities leading to zoonotic disease vulnerabilities seemed primarily related to security challenges. Drivers of migration may cause conflict, in turn causing further displacement and insecurity, negatively affecting ecosystems and nutrition, and through this human and animal immune status. Conflict and insecurity in Nigeria have caused one of the world’s most complex displacement crises, with over 2.7 million people living displaced due to overlapping conflict, disasters, and violence linked to crime (IDMC, 2021). It is essential to acknowledge and better understand the security challenges when addressing zoonotic disease risks in this context. Although the root causes of the violence and insecurity are complex, demanding their own field of study, we highlight some of the grievances of the participants in the below section to support local, contextualised solutions.

Conflicts between pastoralists and farmers are attributed to increased competition over rural resources and land, mixed with local politics along ethno-religious affiliations (Majekodunmi, 2022). Competition has led to the destruction of crops, encroachment of grazing routes and livestock theft, which has increased distrust between crop farmers and pastoralists (Sani Ibrahim et al., 2021). While the expansion of farming ranges hampers access to transhumance routes, which are essential to pastoralists for seasonal grazing, they often get blamed for the destruction of agriculture and crops (Lenshie, Okengwu, Ogbonna, & Ezeibe, 2021; SIPRI, 2022; UNOWAS, 2018). Climate change further exacerbates the risk of farmer–herder conflicts, in areas where governance systems create inequalities (SIPRI, 2022), or where state authority is absent (Lenshie et al., 2021).

Government bans on open grazing negatively affects pastoralist communities. The symbiotic tradition whereby livestock benefited from crop residues in return for manure has collapsed over time, and the fragmentation of land and changes in property rights have further strained relationships between farmers and pastoralists (Sani Ibrahim et al., 2021). Participant interviews identified varied levels of resentment ‘land owners may replant some other forms of crop in the same farmland [as we plant our crops] rendering [our] own crop useless’ (WS2). While many farmers therefore no longer require the traditional services of the Fulani pastoralists, the latter are still dependent on farmers for access to natural resources (Ducrotoy et al., 2018). Our study shows that relations between pastoralists and land-owning farmers are complex, implying that some decisions may be personal rather than practical, which can be explored in finding solutions.

## Conclusion

We found that mobility may mitigate or exacerbate zoonotic disease risk, with the latter in turn leading to forced migration. Security issues such as conflict, rustling and crimes need to be addressed simultaneously with mitigating structural inequalities to reduce disease transmission risk. This includes increasing pastoral social and economic capital to reduce inequality and competition, better access to grazing routes and boreholes, securing land tenure arrangements to reduce the pressure on rangeland resources and minimize conflict, and enhance (veterinary) public health to reduce zoonotic disease risks for the communities and wider population. Importantly, framing of zoonotic disease risk should not blame any specific population group or individuals, to allow for a more inclusive approach to mobility and health.

## Limitations

The qualitative research methodology does not aim to provide generalizable research findings, acknowledging that people and phenomena depend on their context and circumstances, although some of its findings may be applicable to similar groups in comparable situations. [deleted for anonymization].

### Availability of data and materials

The data that support the findings of this study are available on request from the corresponding author. The data are not publicly available due to privacy or ethical restrictions.

## Supporting information

Interview guide

## Declarations

## Acknowledgements

We would like to thank all participants for sharing their time and stories with us, and the local researchers who facilitated the project.

## Ethics

The study protocol was approved by [deleted for anonymization]. Participants were informed about the study during face-to-face recruitment, and informed about the voluntary bases of participation and their right to withdraw prior or during data collection. All participants gave verbal consent for data collection and recording of interviews. Participants’ anonymity was ensured by full anonymization in the analysis and discussion to remove any personal identifiable information.

To minimize the risk of exploitation and damaging research practices among vulnerable and marginalized community participants, we used semi-structured rather than closed questionnaires allowing participants to refocus discussions. [sentence deleted for anonymization].

## Figures and Tables

Figures 1-5 are available from the corresponding author upon request.

**Figure 6:**
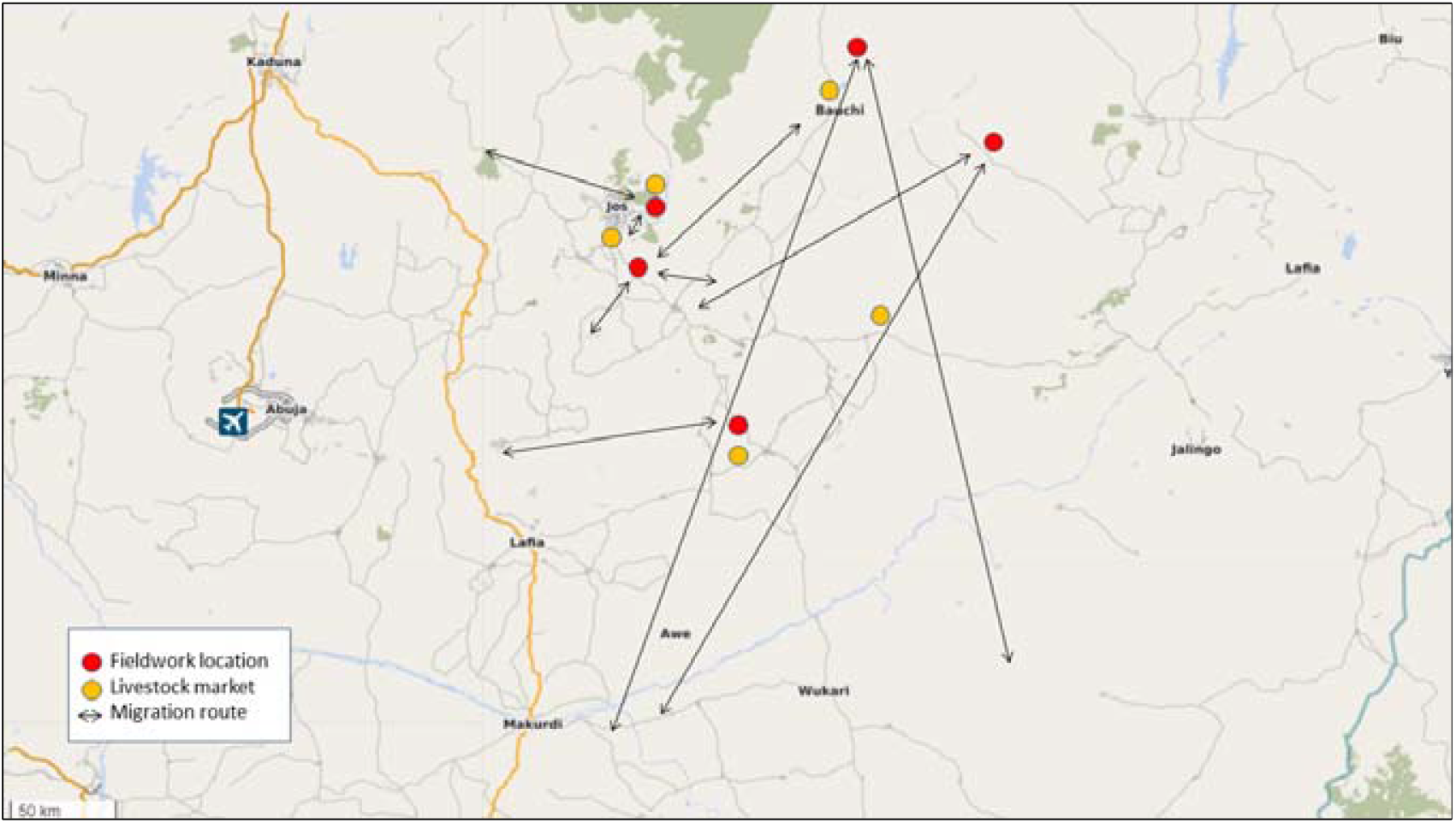
Location of fieldwork and migration routes + market locations (adapted from Open Streetmap)

