## Supplementary material for "A One Health approach to pastoral (im)mobility, health, and disease: a qualitative participatory study in Plateau State, Nigeria": Interview guide

**Livelihoods and livestock**

What are the primary livelihood activities in the household/ community?

If agriculture: what type of crops; where and how are these grown?

If livestock: what are the main animals kept for livelihood purposes?

What are the main animal diseases for: cattle, sheep and goats, poultry?

- can you list the most important diseases and indicate when these occur (seasonal)?

- are you familiar with 'zoonoses', diseases transmissible from animals to humans?

what do you do when an animal falls sick?

What other animals live in the house/ village, for what purpose are these kept?

Do you and/ or your animals interact with wildlife; what are the challenges?

Where are the animals grazed?

Do you have access to a livestock market; how far is the nearest market; if no, how do you market your animal/ products?

What are the challenges in accessing livestock (product) markets?

**Challenges and access to services**

What are the main challenges faced by your community/ household, in terms of livelihoods, access to services, security, etc?

Do you have access to public services, such as education, health?

What are the most important human diseases?

What is your primary water source, how do you get access, and who is in charge of collecting water; do humans and animals access the same water source?

How is waste water dealt with; other garbage; livestock manure?

**Mobility:**

Has anyone in the community, households, individuals migrated;

If yes: what were the main reasons for doing so; permanent or temporarily/ seasonally; with or without animals?

- what are the main challenges during migration?

If no: why did you decide to stay in this location; what are the main reasons for abandoning pastoralist transhumance?

Has your household/ community faced any land ownership challenges?

- How are any challenges resolved; by whom?

**Transect walk:**

- participatory map showing primary health care units, schools, police station, market, river, grazing area;

- discussion on village layout in terms of wealth, tribes;

- walk through community noting: location livestock (market); water points, erosion, etc.
